# Spatial Point Process Model Predicts Survival in Clear Cell Renal Cell Carcinoma

**DOI:** 10.1101/2022.02.16.22271019

**Authors:** Prahlad Bhat, Michael Hwang, Afrooz Jahedi, Kanishka Sircar, Kasthuri Kannan

**Affiliations:** Department of Translational Molecular Pathology, UT MD Anderson Cancer Center, Houston, Texas; Department of Pathology, Indiana University, Indianapolis, Indiana; Department of Pathology, UT MD Anderson Cancer Center, Houston, Texas

**Keywords:** clear cell renal cell carcinoma, sarcomatoid, spatial point pattern, pair correlation function, Gibbs process

## Abstract

Among renal cell carcinomas (RCCs), clear cell RCCs (ccRCCs) is a highly aggressive class characterized by highly invasive clinical course and poor survival. The presence of a sarcomatoid component implicates an even poorer prognosis. Spatial measures can establish features that could be routinely used in clinical practice to stratify important RCC outcomes. Therefore, we tested the effectiveness of spatial features as predictors of survival differentiation in 58 Grade 2 ccRCCs. We developed a machine learning model using the acquired spatial features to predict survival in Grade 2 ccRCCs which we termed “SurvCal”. The receiver operating characteristic accuracy from the derived model was 0.812. Subsequent feature analysis identified the spatial model fitting intensity parameter Gamma and the pair correlation function as critical features in distinguishing the classes. In the light of the increasing digitization of pathology routines, our results demonstrate the importance of spatial point pattern features as determinants of ccRCC survival outcomes and phenotypes.

## 1. Introduction

Among renal cell carcinomas (RCCs), clear cell RCCs (ccRCCs) are the most common, accounting for 70-80% of all RCCs [1]. Owing to the increased use of imaging studies, the detection and incidence of RCC have risen steadily over the past few decades. However, there has not been a corresponding rise in RCC mortality, as many incidentally detected RCCs are indolent [2]. Differentiating aggressive RCC from indolent RCC through prognostic biomarkers remains a significant challenge that directly impacts patient management. A prognostic marker for an aggressive subtype of RCC is the sarcomatoid differentiation in RCC (or sRCC), which accounts for only 5% of all RCCs [5] but comprising of approximately 20% of metastatic RCCs [6]. In sRCC, some tumor cells lose their epithelioid morphology and acquire a mesenchymal sarcoma-like morphology. RCCs comprise varying amounts of sarcomatoid cells. However, any small fraction suffices the RCC to be considered an sRCC and portends a poor prognosis [7]. Tumor grade is independently prognostic, mainly as prognostic nomograms for grades 3 and 4 RCCs [3,4], and sRCCs constitute grade 4 RCCs.

Tumor grading using preoperative renal mass biopsy is inaccurate given the heterogeneity of different RCC grades and the limitations of examining only a small portion of the tumor that may not include the highest-grade components. Identifying sarcomatoid features suffers from this drawback since surgical biopsy identifies sarcomatoid morphology in less than 10% of sRCC cases [8]. Moreover, mere visual inspection of epithelial ccRCC tissues cannot detect differences in the nucleic morphometry that may indicate sarcomatoid dedifferentiation elsewhere in the tumor. Therefore, computational schemes to identify sarcomatoid dedifferentiation preoperatively by sampling the epithelioid ccRCC component will improve patient stratification and management as biomarkers for sRCCs are yet to be discovered.

Because sarcomatoid dedifferentiation involves dynamic cell mobilization and rearrangement, the main differences between ccRCCs and epithelioid sRCCs lie in the difficulty of visualizing the spatial organization of cells rather than more easily discernable nuclear morphometry. The mutations driving ccRCC and sRCC tumorigenesis are distinct [9], possibly reflecting the tumor phenotype. Therefore, differences in the spatial organization of cells in the tumor microenvironment (TME) of epithelioid sRCCs and ccRCCs may reflect differences in these tumors’ biology. Using spatially quantifiable measures that can assess patterns invisible to the naked eye, we can significantly improve the clinical predictability of sarcomatoid dedifferentiation over traditional qualitative and morphology-based classification.

Similarly, ccRCCs associated with long-term (LT) and short-term (ST) survival in patients, particularly low-grade ccRCCs, share many morphological characteristics. Because low-grade ccRCCs are a heterogeneous group, with some cases still in the initial stages of tumor progression, visual inspection to predict the prognosis of these tumors is challenging. Retrospective studies have shown that although RCC patients with grade 1 or 3 tumors have distinct survival curves, those with grade 2 or 3 tumors have overlapping survival curves that do not delineate by the World Health Organization/International Society of Urologic Pathologists grading. Given that grade 1 RCC is exceedingly rare and that the bulk of RCC tumors is grade 2 or 3 [10], survival is only reasonably predicted with overall nuclear morphology, with much room for improvement. Because infiltrative processes mediate by coordinated rearrangements of the cells’ distributions, which are vital to tumor progression, it is imperative to incorporate spatial characteristics of the TME as a biological predictor of survival outcomes.

Therefore, we tested the effectiveness of spatial point pattern measurements as predictors of survival in 58 ccRCCs images with 3 annotations per image. To generate spatial measures useful for biological inference, we either need to estimate parameters from fitting spatial models or derive spatial statistics. In this project, we did both: we estimated model parameters by fitting a derivative of a spatial inhibition model – a Gibbs process, called the Strauss process. and computed a spatial statistic called the pair-correlation function. In addition, we computed morphological metrics such as nucleus size, mean cell cellularity etc. We refer to these quantities - the parameters from model fitting, the pair correlation function statistics, and morphological metrics collectively as “spatial features.” We developed a machine learning model using the acquired spatial features to predict survival in Grade 2 tumors : SurvCal. For the SurvCal, the receiver operating characteristic (ROC) accuracy was 0.812. We identified the Gamma index and pair correlation function as critical features distinguishing the classes for these classifiers. In what follows, we describe the methods and results in detail, followed by a discussion.

## 2. Materials and Methods

### 2.1 Sample Collection

The MD Anderson-derived tissues were processed via standard Formalin-Fixed Parrafin-Embedded techniques as illustrated by Sadeghipour et al. [13] and imaged. We derived whole slide images of the tissues from three repositories. Most tissues came from the TCGA GDC portal, where we queried for diagnostic tissue slides. Tissues were selected irrespective of age, sex, or stage of tumor of patients and were blocked solely on grade. We selected one type of tissue – Grade 2 ccRCCs using two distinct methods – Microarray blocking and. For SurvCal, we collected grade 2 ccRCCs, labeled as G2-ccRCC. Table 1 summarizes the tissue information and the dataset sources.

**Table 1:** Tissue and Data Sources Information

| Description | G2-ccRCC (58) |
| --- | --- |
| No. of tissues in the training set | 44 |
| No. of tissues in the external validation set | 14 |
| Total no. of annotations<br>(n=3 per tissue) | 174 |
| Fuhrman grade 2 | Yes |
| Fuhrman grade 3 | No |
| Fuhrman grade 4 | No |
| No. of tissues from MDACC microarray repository | 8 |
| No. of tissues from MDACC diagnostic tissue repository | 0 |
| No. of tissues from TCGA | 50 |

### 2.2 Analysis workflow

The analysis workflow consists of three steps: 1) obtaining morphological features and generating spatial point pattern objects, 2) obtaining spatial statistics and model fitting parameters, and 3) classification and feature selection. Figure 1 illustrates this workflow.

**Fig. 1.**
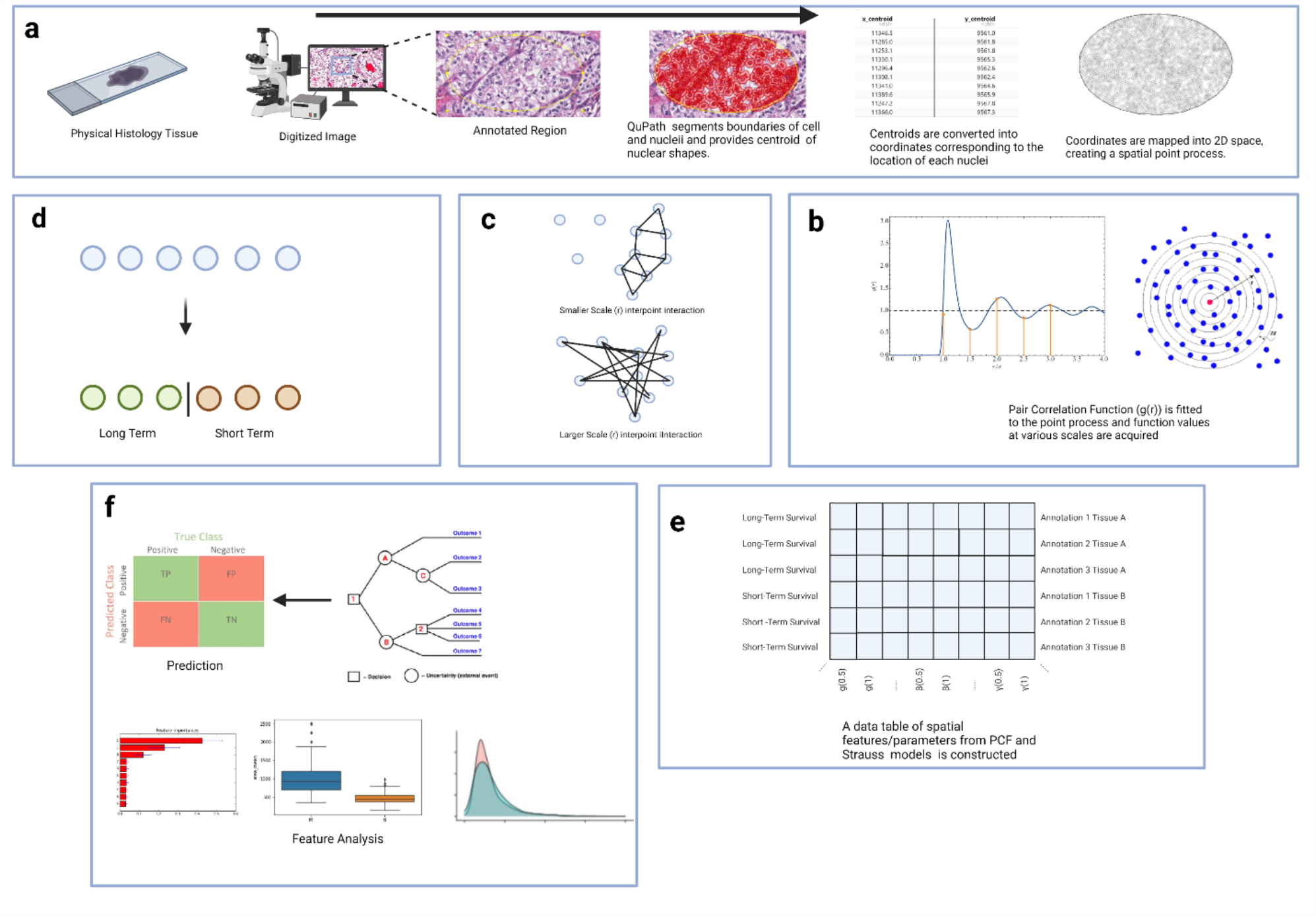
Analysis workflow. (a) Physical histology tissues are derived from the surgical resection of tumor masses in the kidney. Tissues are then digitized, and pathologists select a small portion of the tumor to be annotated. The annotations are then segmented into cell and nuclei components. The segmented nuclear regions’ barycenter coordinates are then calculated and converted into a spatial object. (b) The Pair Correlation Function and Strauss model is fitted to each annotated tissue region and spatial measures are derived. (c) The concept of the scale of interaction in point processes. Functions that are computed over large scales measure interactions between cells that are more distant from one another, whereas functions that are computed over small scales measure interactions between cells that are nearer to one another.(d) Tissues are stratified into groups based on survival time in weeks.(f) Spatial Features derived using Pair Correlation Function and Strauss models are inputted into a machine learning classifier to predict survival group. Additionally, spatial differences between ST and LT groups are analyzed.

#### 2.2.1 Obtaining features and generating spatial point pattern object from digitized Whole Slide Images (WSI)

WSI was loaded into QuPath [14], a pathology suite for viewing and processing whole slides, for a pathologist’s review. The pathologist determined three regions per tissue and annotated them, corresponding to the highest Fuhrman grade. We selected the tissue regions closer to tumor cores rather than leading edges to avoid variation in nucleic densities. The annotations varied in size but were usually no larger than 1000 × 1000 µm^2^. We avoided regions such as stromal infiltration, blood vessels, and immune cells. QuPath’s segmentation algorithms segmented the annotated tissue regions, and the cell-detection algorithm identified the coordinates of the nucleus center in each cell. We exported these features to generate the spatial point pattern object for model fitting and spatial statistics. Figure 1 highlights the workflow followed for this step in the analysis workflow.

#### 2.2.2 Acquiring fitting parameters and spatial statistics

Once the locations of nuclei in each annotated tissue space were extracted, spatial estimates for each annotation were derived that quantified cell interactions. The method used to derive spatial features was the conversion of nuclei locations within the annotation into a Strauss process by first fitting an appropriate spatial model and a subsequent application of a spatial function on the process.

A spatial point process is a bounded collection of points in a defined space. This is the basic framework from which all the spatial data can be collected. Using these point processes, a Pair Correlation Function (PCF) was applied to obtain the statistics. Increasing radial values represented an increasing scope of interaction while corresponding function estimates showed the degree of interaction at a specific scale (Fig. 1b). The PCF was computed along a non-continuous interval of [0 *µm*, 50 *µm*] with increments of 0.5 *µm*. This led to 101 radial distances being applied as inputs for the PCF. In addition, an Akaike information criterion (AIC) was obtained for each point pattern as a summary statistic to fit an optimal Strauss process model to each annotation. This was done by selecting the fitted Strauss model that corresponded to the highest AIC statistic for each point pattern.

These computations resulted in 101 spatial predictor variables per annotation in addition to an optimal *r* and Gamma values (from the optimized Strauss models).

#### 2.2.3 Patient Stratification using K-Means Analysis

Because a machine learning approach was utilized in this study to predict patient survival, grouping the G2 ccRCC tissues into “classes” based on the survival for each patient provided a simple framework for classification and comparison, which Fig. 2 illustrates. Determining the optimal number of groups was conducted by calculating the number of groups such that intragroup variation is minimized using the sum-of-squares method (), as illustrated in Fig (2a).Two-means clustering determined that the mean survival time that separated the groups was 1594.5 days (Fig.2b). All ccRCC samples above and below this mean were denoted as Long-and Short-Term Survivors - “LT” and “ST” respectively.

**Fig. 2.**
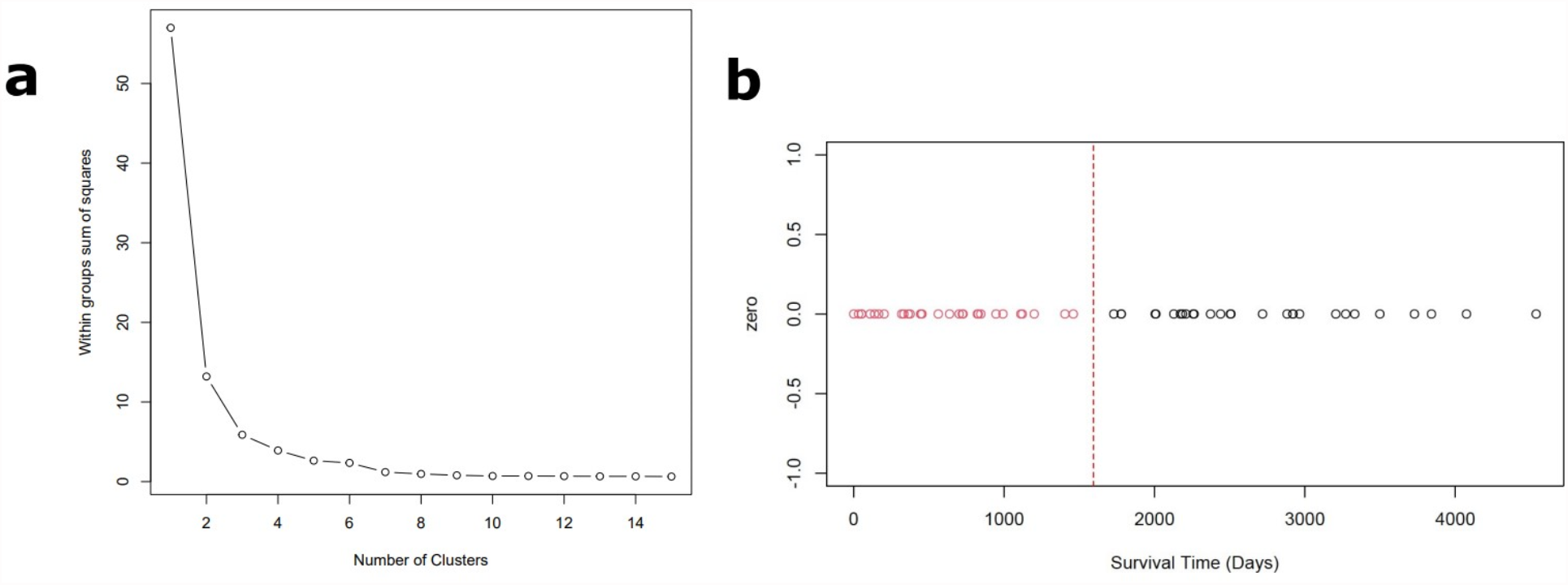
K-means Analysis. Fifty-eight grade 2 tissues were selected at random from MD Anderson microarray repositories and TCGA repositories, and a 1-dimensional k-means analysis was applied to the sample, with survival time as the clustered variable. (A) Greatest drop-off in sum of squares variation shows that two clusters is optimal for analysis. (B)The cluster analysis showed that tumors associated with survival times below and above 1600 days constituted 2 distinct groups.

#### 2.2.4 Machine Learning and Feature Analysis

In this study, spatial features were used to predict survival in G2 ccRCC patients using the eXtreme Gradient Boosting (XGBoost) R package, a high-performance machine learning architecture. The model was was trained and tested on a randomly selected validation cohort (44-14 train-validation split) from the existing G2-ccRCC dataset. The features used in the training model were the optimized r and gamma parameters of the optimized Strauss Process corresponding to each ccRCC tissue region, as well as PCF values of each point process from intervals of [0,50] selected from our entire tissue. After model building, the distributions of these spatial features were compared between the LT and ST groups defined in Fig 2.

## 3. Results

### 3.1 Survival is Associated with Spatial Morphology

In a Strauss Model, an R parameter describes the interaction distance. Larger r values can be interpreted as cells being more dispersed from each other than in spatial models with smaller r values. As shown in Fig 3a, ST G2 ccRCCs tend to have Strauss models with larger median r parameter value than LT tumors (LT = 6.8 *um*, ST = 7 *um* p = 0.0079). Thus, Tumors with fairer prognosis have cells that are farther apart from one another, which would suggest dispersive tendencies in tumors within patients with longer survival times. This is further corroborated by the Gamma parameter in Fig 3b., which illustrates that ST tumors have higher median gamma parameters from the fitted Strauss models than LT tumors do (LT = 0.053, ST = p = 0.069). Both LT and ST G2-ccRCC tissues have fitted Strauss models with gamma parameters < 1, so it can be concluded that G2 ccRCC tumors overall tend to have cell spacing that behaves in a clustered pattern but ST tumors cells tend to be more ordered relative to LT tumors. The PCF distributions for LT and ST groups reveal a higher proportion of larger PCF values for ST tumors and a larger proportion of PCF values centered around 1 for LT Tumors. This illustrates that ST tumors tend to display cells with higher degrees of infiltration, a hallmark mechanism of cell migration and metastasis which dramatically reduce lifespans in ccRCC patients.

**Fig. 3.**
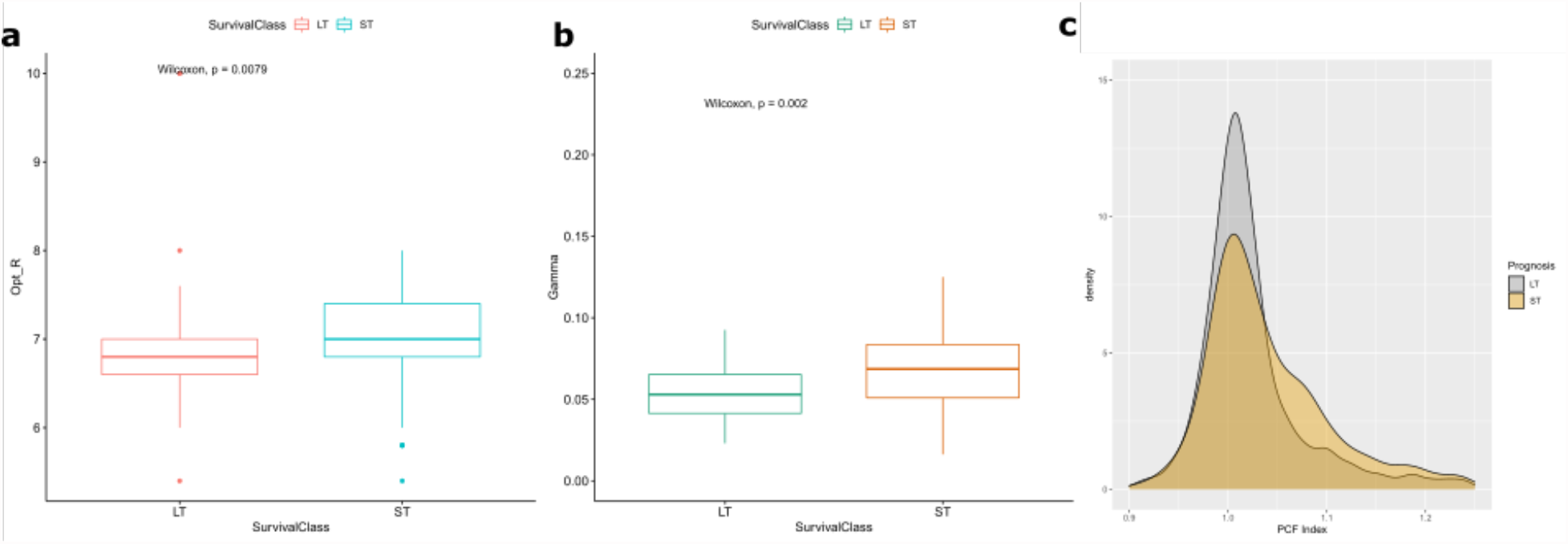
Spatial Feature Comparisons. The G2-ccRCC tissue set was split along the survival classes defined as “LT” and “ST”. The tissue count refers to the number of annotations. Fig 3a. illustrates the overall differences between the Strauss Process-derived optimal R distance in LT and ST groups. Fig 3b. illustrates the overall differences between the Strauss Process-derived optimal Gamma value in LT and ST groups. Fig 3c. Compares the distribution of PCF values of all G2-ccRCC tissues from [0,50] between ST and LT tumor type.

ST tumors display aggressive cell behavior, which supports the idea that subtle differences in tumor microenvironments of ccRCCs can alter prognosis. The differences between Gamma and Opt_R parameters in LT and ST tumors constitute that are impossible to discern with pathological visual inspection alone. However, there are significant differences that can be quantified by spatial modelling. ST tumors contain cells which scatter more readily but display more significant inhibitive tendencies while also still being more infiltrative than LT tumors. This could suggest that infiltration in G2 ccRCCs also entails an organized and progressive change in cell-cell contact.

### 3.2 Modelling using XGboost Proves Clinical Robustness of Spatial Features

Using the XGBoost trained model SarPCal, we were able to achieve clinically significant model accuracy (AUC = 0.8798)(Fig. 4a) for predicting survival outcomes in grade 2 ccRCCs, respectively. Each fold accuracy in the model’s cross-validation is shown in Fig. 4b. This model elucidates the effectiveness of spatial features derived from model building and statistical metrics as clinical predictors of the ST and LT groups. However, the external validation AUCs for the SarPCal classifier outperformed the results of the mean cross-validation AUCs, indicating model robustness. This may have been due to the fact that the method of data collection for the training model resulted in the inclusion of tumor regions from different parts of whole diagnostic tissues that were vastly heterogenous morphologically, allowing for a more stable model.

**Fig. 4.**
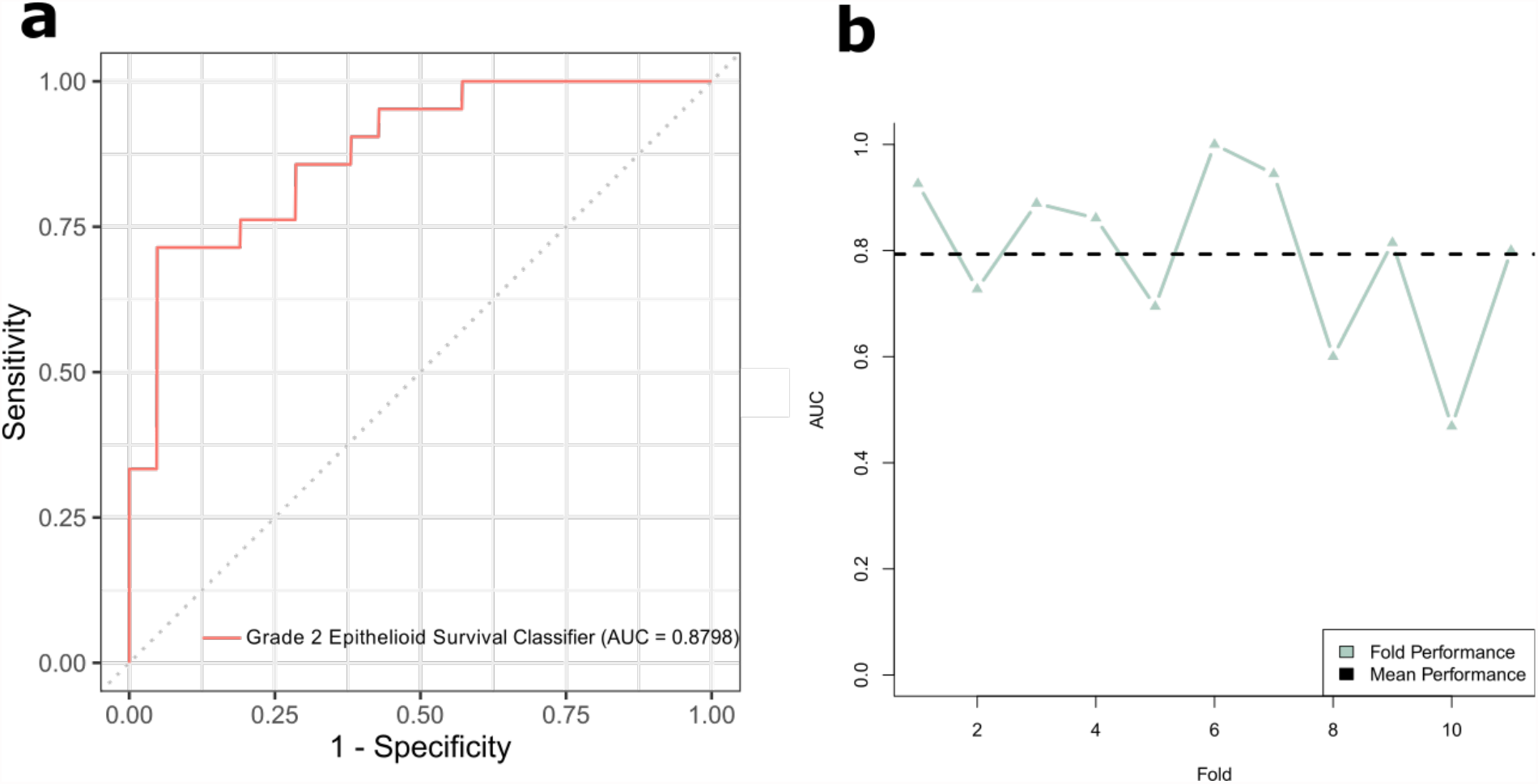
Survival Model Evaluation. XGBoost model SarPCal was evaluated on G2-ccRCC dataset. Fig 4a. shows the Receiver Operating Curve (ROC) of an external validation cohort (n=14) AUC against the training cohort (n=44). Fig 4b. shows the cross-validation AUC for each training fold using 11-fold validation on the training cohort. Large variations in cross-validation AUC between folds suggests high model robustness.

## 4. Discussion

This was the first study to use quantitative spatial features to distinguish sarcomatoid tumors from non-sarcomatoid tumors using only the epithelioid components of the tumors. Moreover, this study was the first to predict the survival outcomes of patients with low-grade ccRCC. The study used spatial features derived from models that utilize cell-cell pairwise distances as metrics for interactive behavior. We built a machine learning model to classify tumors as sarcomatoid dedifferentiated or non-sarcomatoid dedifferentiated tumors as well as poor- or good-prognosis tumors. This study demonstrated the effectiveness of using spatial features to predict tumor groups and the promise of using machine learning architectures to derive meaning from spatial statistics. RCC grading, which is based almost exclusively on the nuclear features of tumor cells, is a morphology-based diagnostic feature that pathologists use to predict survival. In one study evaluating the efficacy of the Fuhrman and World Health Organization/International Society of Urologic Pathologists grading methods, both grading systems were shown to be in moderate agreement with observers’ assessments (kappa=0.34 and 0.48, respectively). This discrepancy indicates that traditional nuclear grading has some level of subjectivity that can be improved with the application of more precise quantitative methods. RCC grading based on preoperative biopsy samples is also inaccurate, having only 50-75% concordance with the final resection grading [16], [17], as upgrading at surgical resection frequently occurs ([18]). Because ccRCCs invariably move, diagnostic predictors should reflect these dynamic spatial interactions among the large majority of tumor cells. Diagnostic predictors should not be limited to considering only a select subpopulation of the most morphologically atypical neoplastic cells. Our study revealed that differences in tumor outcomes are more readily defined by cellular infiltration than by pairwise cell-cell interactions such as repulsion and attraction. Cells undergoing epithelial-mesenchymal transition can alter their extracellular matrix, lose apical-basilar polarity, and lose contact inhibition to facilitate local invasion; thus, from a spatial perspective, cells lose many of the inhibitive and interactive proper-ties they possessed prior to differentiation. Our study showed that tumors associated with a fair prognosis had more spatially organized, clustered cell bodies than did tumors associated with a poor prognosis. These results are supported by the fact that aggressive tumors become rapidly disorganized as they progress, resulting in highly unusual organizational patterns.

Despite the morphologic heterogeneity of sRCC, its epithelioid and sarcomatoid components both have a gene expression signature indicative of poor prognosis. The global gene expression differences between non-sRCC and sRCC are greater than those between the epithelioid and sarcomatoid components of sRCC [19] [20]. These findings suggest that genomic aberrations can cause spatial phenotypic alterations that are reflected by quantifiable spatial measures of cell behavior. This study was the first to quantifiably prove the morphological distinction between epithelioid sRCC and non-sarcomatoid ccRCC. Compared with non-sarcomatoid ccRCCs, epithelioid sarcomatoid ccRCCs are less infil-trative and have more varied densities. This is unexpected, because sRCCs tend to be much more aggressive than ccRCCs, even at grade 4. This finding is partially supported by evidence that suggests that sarcomatoid and non-sarcomatoid ccRCCs have different progenitors and driver mutations, which could affect the tumor’s phenotypes and thus the spatial distributions and organization of their cells.

Previous studies that attempted to use quantifiable morphology-based features to stratify disease outcomes or types relied mostly on cellular and nuclear measurements, such as area, perimeter, and roundness. Although these features have clinical implications, they do not reflect omnipresent dynamic cell interactions. Whereas morphometric features can be visually assessed, spatial arrangements cannot. Our pathological spatial classifier combined morphology- based features with spatial metrics to generate a highly accurate model. The classifiers built in this study can maintain high sensitivity with clinical application, thereby providing pathologists with a novel predictive tool for risk-stratifying patients with grade 2 ccRCC and preoperatively identifying sRCC from just the epithelioid component, without the need to sample the sarcomatoid component. This study was not without its limitations. The tissues we analyzed were taken from multiple regions from several tumors; however, the tumor region demarcation method we used involved selecting regions with minimal stromal infiltration. This design was somewhat biased and may have yielded samples that were not entirely representative of the whole tumor. This was partially addressed by our study’s large sample size and large annotation feature col-lection. We captured basic morphometry of nuclei as predictors to improve model predictability, but we did not calculate any measures of spread such as median, standard deviation, kurtosis, and skewness of these features, which limited the number of features that the XGBoost models in this study had to work with. Additionally, many of the mathematical methods used to quantify cell-cell interactions or dispersive tendencies were overly broad in scope, and further analysis that correlates spatial gene expression patterns to phenotypic alterations could elucidate changes in cell dynamics in ccRCCs. However, this study provides conclusive proof that spatial point theory holds great promise in understanding invisible cell dynamics and their correlation to clinical outcomes in ccRCCs.

In summary, we demonstrated that spatial histopathology measures of cellu-lar interaction can predict tumor outcomes such as prognosis and sarcomatoid dedifferentiation. We effectively established predictive models and used feature differences to classify tumors by their predicted outcomes. We also discovered distinct modalities of cell locomotion in ccRCC. Our methods can be repro-duced in a clinical setting with HE slides, in which the spatial characteristics of tumors can reveal underlying biological processes and guide pathologists to make diagnoses that are more accurate than those achieved with contemporary brightfield microscopy. The use of spatial point processes to model the TME should be applied in future studies of other epithelial cancers to assess these processes’ broad clinical applicability.

## Data Availability

All data produced in the present study are available upon reasonable request to the authors

